# Intra-tumoral epigenetic heterogeneity and aberrant molecular clocks in hepatocellular carcinoma

**DOI:** 10.1101/2021.03.22.21253654

**Authors:** Paula Restrepo, Adrian Bubie, Amanda J. Craig, Daniel Cameron, Ismail Labgaa, Myron Schwartz, Swan Thung, Gustavo A. Stolovitzky, Bojan Losic, Augusto Villanueva

## Abstract

There is limited understanding of the epigenetic drivers of tumor evolution in hepatocellular carcinoma (HCC). Here we characterize the epigenetic contribution of methylation to intra-tumoral heterogeneity (mITH) using regional enhanced reduced-representation bisulfite sequencing DNA methylation data from 47 early stage, treatment-naive HCC biopsies across 9 patients by quantifying regional differential methylation across promoters and CpG islands, while overlapping with methylation age markers. Furthermore, we integrate these data with matching RNA-sequencing, targeted DNA sequencing, tumor-infiltrating lymphocyte (TIL), and hepatitis-B viral expression data. We found substantial mITH signatures in promoter and enhancer sites across 44% of patients in our cohort that highlight a novel axis of ITH that is not otherwise detectable from RNA analysis alone. Additionally, we identify an epigenetic tumoral aging measure that reflects a complex tumor fitness phenotype as a potential proxy for tumor clonality. Associating clinical outcomes with epigenetic tumoral age using 450k array data from 377 patients with HCC in the TCGA-LIHC single-biopsy cohort we found evidence implying that epigenetically old tumors have lower fitness yet higher TIL burden. Our data reveal a novel, unique epigenetic axis of ITH in HCC that merits further exploration.

## INTRODUCTION

Hepatocellular carcinoma (HCC) is a common yet lethal form of liver cancer that has one of the fastest growing incidence rates globally,^1,2^ typically arising in patients with a background of chronic liver disease, such as hepatitis B or C (HBV or HCV) infection, alcohol use disorder, and metabolic dysfunction associated steatotic liver disease (MASLD). Despite recent improvements in systemic therapies, drug resistance remains a challenge and median survival in advanced stages is still less than two years. We and others have shown significant molecular intra-tumoral heterogeneity (ITH) in 30-40% of HCC.^3–5^ This includes complex mixtures of subclonal cancer populations evolving over time and interacting with non-tumoral stromal and immune cells in the microenvironment,^6^ a key molecular feature underlying drug resistance in HCC.

Previous studies of ITH in HCC have leveraged experimental designs with multi-regional sampling from the same tumor.^3,4,6–8^ These studies found that driver mutations in genes such as *TERT* promoter, *TP53*, and *CTNNB1* tend to be founder events present in all regions of the tumor, and increased ITH correlates with worse prognosis and clinical outcomes in patients with HCC.^3,4,7,8^ Our previous work found distinct ITH-dependent patterns of spatio-temporal interactions between HCC and immune cells along with strong evidence of spatially heterogeneous immunoediting in multi-regional samples,^3^ indicating that HCC ITH cannot be fully characterized through mutation data alone.^3,4^

One notable yet relatively underexplored aspect of ITH in HCC is the role of DNA methylation. Previous examination of epigenetic drivers highlight promoter hypermethylation of tumor suppressor genes as an important mechanism for tumorigenesis and disease progression in HCC and some of its background etiologies.^9,10^ We detected aberrant methylation patterns in HCC that can be distilled into signatures predictive of patient prognosis and survival, indicating a potentially useful avenue for biomarker discovery.^9^ A previous study examining DNA methylation using multi-regional sampling in a small cohort of HCCs reported significant epigenetic heterogeneity among tumors, in contrast to strong interindividual homogeneity of nonmalignant liver tissue.^4^

A key aspect of methylation data is its ability to predict molecular epigenetic age, which measures proxies of biological age of a tissue as opposed to its chronological age.^11^ These “epigenetic clocks” have been extensively validated in normal tissue and used to examine the biological aging process and its relation to human diseases, including cancer.^11–13^ However, the role of methylation age in ITH has not been well characterized in HCC to date.

To further investigate the unique contributions of DNA methylation to HCC evolution and ITH, here we use enhanced reduced representation bisulfite sequencing (eRRBS) to obtain methylation profiles from multiregional biopsies of 9 previously analyzed patients.^3^ By adding another layer of methylation data to existing molecular ITH profiles, we explicitly compare ITH signatures derived from RNA expression and DNA methylation data and find key differences. We leverage well-known associations between conserved methylation signatures and chronological patient age to derive estimates of differentially aging tumoral regions that reflect novel aspects of tumor evolution and heterogeneity not captured by expression profiling alone.^11,12^ Integrating the TCGA-LIHC (HCC) cohort with our data, we find a differential tumoral aging profile that strongly associates epigenetically younger TCGA-LIHC tumors with greater DNA clonal diversity, tumor mutation burden (TMB), tumor fitness (poorer patient prognosis), and lower tumor infiltrating lymphocyte (TIL) burden. The differential regional aging of tumors is a novel axis of epigenetic ITH which may reflect a complex tumor fitness phenotype, serving as a useful proxy for tumoral evolution and ultimately the prognostic stratification of patients.

## RESULTS

### Spatial tumor methylation profiling via multi-regional sampling

We performed enhanced reduced representation bisulfite sequencing (eRRBS)^14^ on 47 samples from 9 HCC patients, including tumor (35 samples, including a technical replicate) and adjacent non-tumoral regions (12 samples, **Fig 1A**). On average, we analyzed a median of 5 samples per patient. All patients were treatment-naive at the time of the surgical resection. Most patients were male (67%) with a median age of 61 years, median tumor size of 6.5 cm, and about 55% (5/9) had hepatitis B infection as the most common underlying liver disease (**Fig 1B**). Drawing on previously reported data for these samples,^3^ tumor purity was determined using SNP-array data via ASCAT (version 2.4) and histological examination.

**Figure 1.**
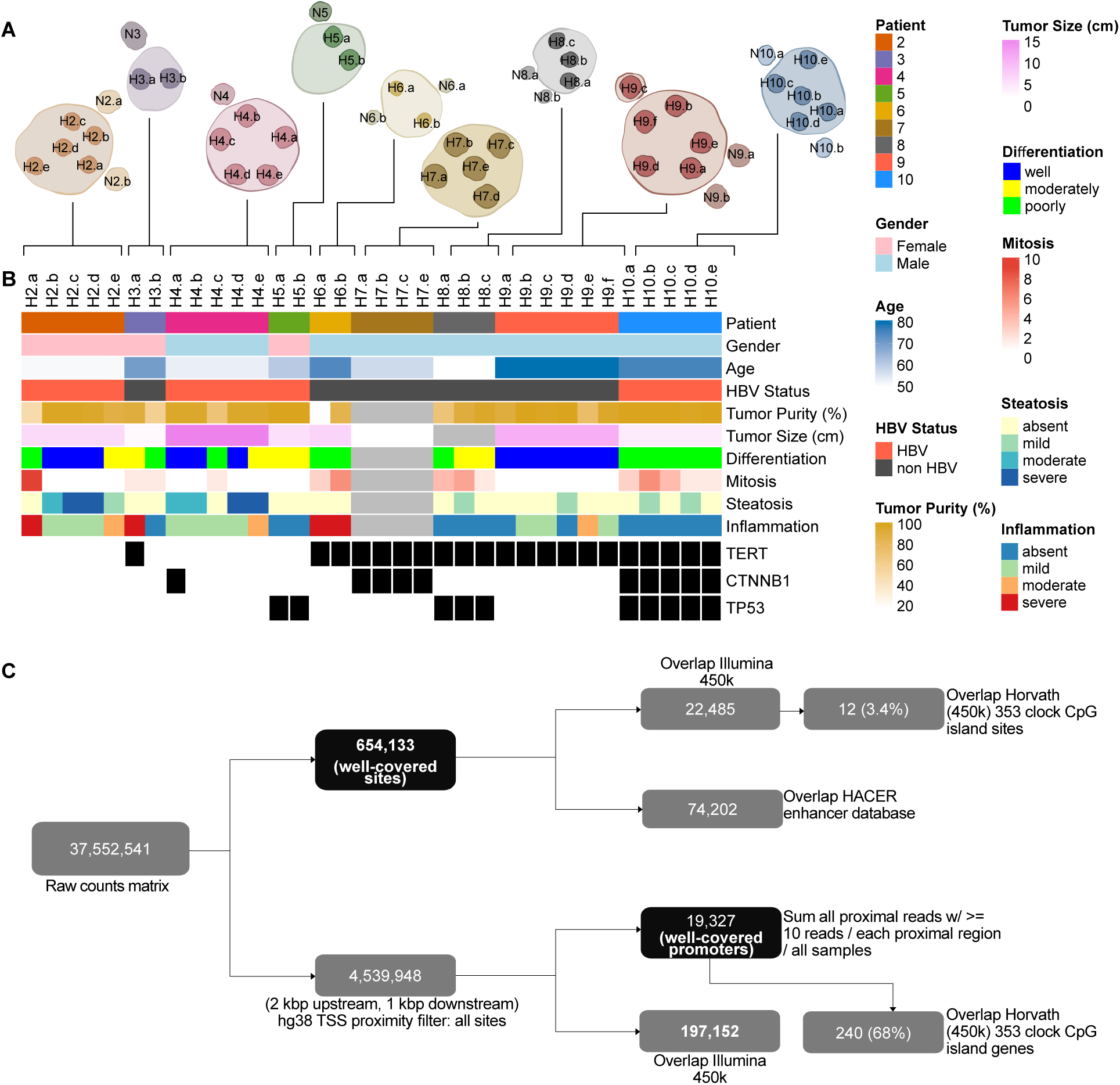
Study cohort and RBBS analysis workflow. A) Representation of multi-regional sampling for tumors used in this study, and available data types used. B) Heatmap showing clinical features of samples, including TERT, CTNNB1, and TP53 mutation status. C) Flow chart indicating RRBS processing and analysis framework.

Since eRRBS data offers a granular view of methylation status at specific CpG sites both within and outside of promoter regions, we designed our regional methylation analysis to take advantage of both promoter-specific and site-specific sensitivity. We performed a site-specific analysis considering only well-covered CpG sites regardless of proximity to a transcriptional start site (TSS). We then aggregated sites within some proximal region of a TSS and conducted essentially a gene-wise promoter-based analysis allowing a reasonable comparison to both 450k methylation array data (and methylation aging signatures derived from it) and bulk RNA-seq transcriptional profiling (**Fig. 1C**).

### Regional epigenetic ITH (mITH) reveals novel differential methylation associations

Using the normalized site-specific methylation data, we performed a PCA analysis to quantify the leading axes of variation driving patient and regional specific DNA methylation signatures (**Fig. 2A**). The first two principal components collectively explain 26% of the total variance, but the leading principal component separates tumor from adjacent normal samples and broadly stratifies tumors by degree of tumor-infiltrating-lymphocyte (TIL) burden (defined by VDJ read burden normalized for library size from RNAseq) (Spearman rho = 0.76, P < 2.2e-16) and number of VDJ clones (Spearman rho = 0.82, P < 2.2e-12) (**Fig 2C**). While there is strong evidence of patient-specific clustering, there are also outlier regions which demonstrate that the relative scale of ITH can overwhelm patient-specific variance, as demonstrated by the second principal component which reveals that patient 8 is a strong outlier (**Fig 2A**), and these conclusions are maintained using the promoter-specific methylation data as well (**Supp Fig 1A**).

**Figure 2.**
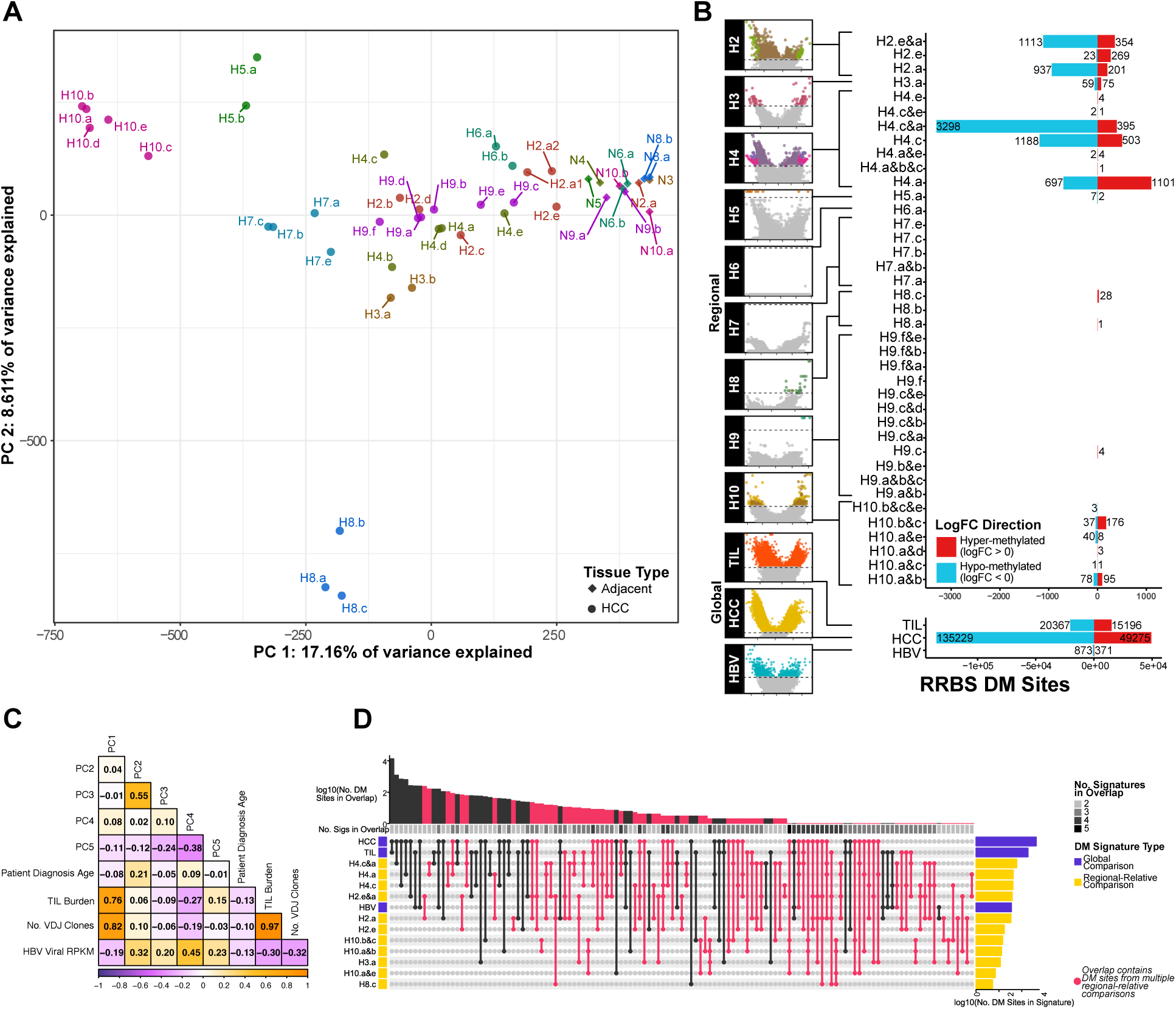
Differentially methylated sites. A) PCA of beta values. B) DM Sites in regional-relative and global comparisons. C) Correlation heatmap of MSSM cohort sample clinical, phenotype, and PCA. D) UpSet plot comparing shared significant DM sites across intra-patient, inter-patient, and global comparisons.

To directly derive multi-regional signatures reflecting epigenetic ITH (mITH) across patients’ tumors, we carried out within-patient differential methylation analyses at both the site (**Fig 2B, Supp Table 1**) and the promoter level (**Fig 3A, Supp Table 3**). Patients 2, 3, and 10 displayed significant mITH (**Fig 2B**, **Fig 3A**), while patient 4 revealed the highest ITH with region H4.c having the highest relative number of differentially methylated promoters (1071 differentially methylated (DM) promoters, FDR < 0.05). Patient 6, previously described as being heavily immune infiltrated displayed only comparatively minor mITH DM at region H6.a (1 DM promoter, FDR < 0.05, **Fig 3A**) despite TCR sequencing showing extensive heterogeneity in TCR across the different tumor regions.^3^ Although patients with more biopsies sampled per tumor have more power to detect mITH, the most densely sampled patient, P9 (n=6), had very few mITH DM sites or promoters, in contrast to patient 3 with only 2 tumor sites sampled, yet detected more heterogeneity (**Fig 2B**, **Fig 3A**). This suggests that our average sampling rate of 5 biopsies per tumor was sufficient to capture the leading effect of mITH.

**Figure 3.**
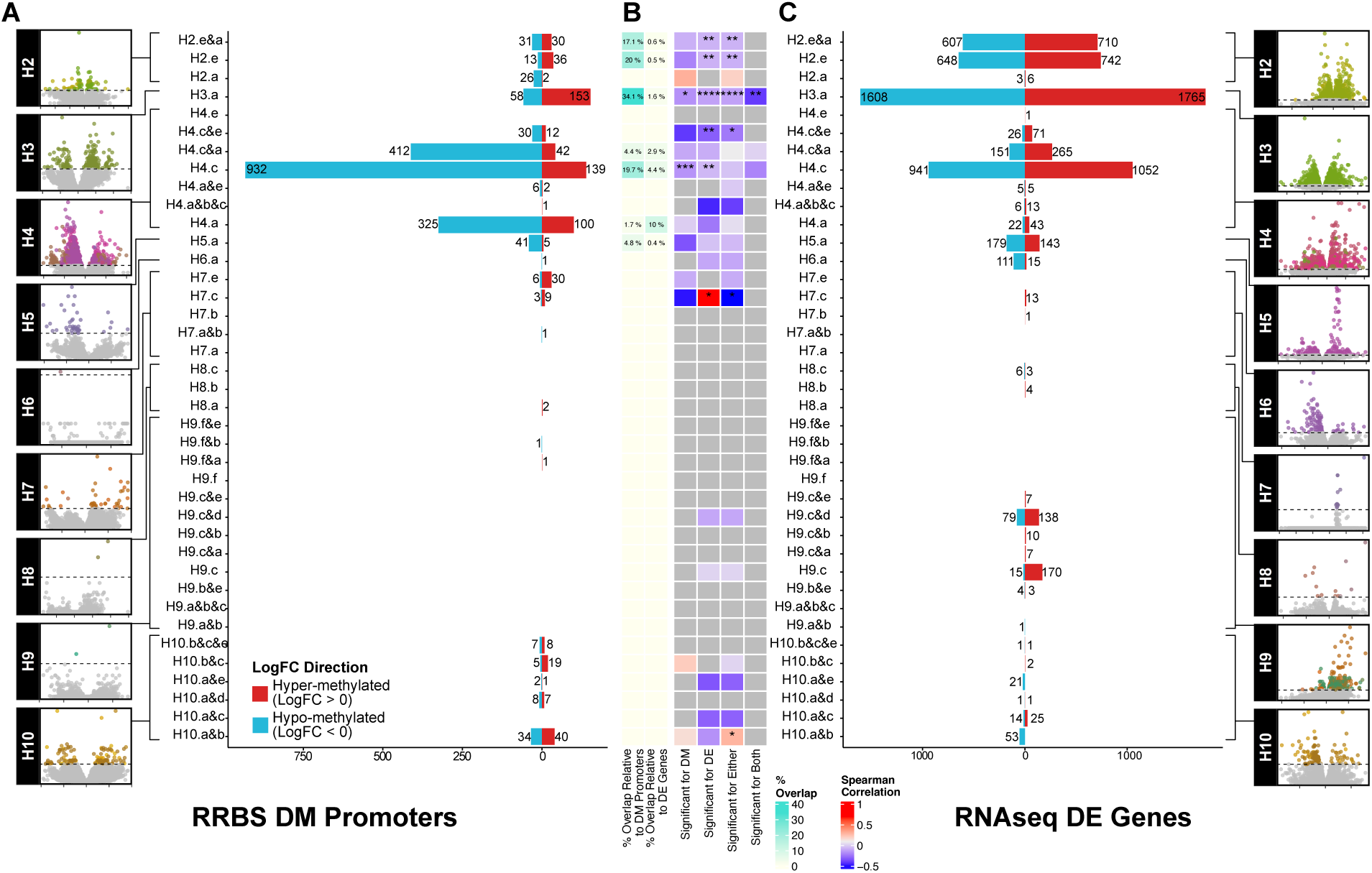
Regional analysis of differential methylation and expression. A) Regional-relative DM promoter profiling. B) Overlap between significant DM promoters from RRBS promoter regions and significant DE genes in matched RNA seq. C) Regional DE gene profiling via RNA-seq.

To compare regional mITH signatures with those associated with HBV viral expression, TIL burden, and tumor vs adjacent normal, we leveraged our previously reported results using these patients.^3^ We identified site and promoter level mITH DM signatures driven by TIL, tissue type (normal vs HCC tumor), and HBV viral expression (**Fig 2B, Supp Fig 1B, Supp Table 2 (sites), Supp Table 4 (promoters)**). As expected, the HCC tumor-specific signature had the most DM sites (FDR < 0.05, 184,504 Sites, **Fig 2B**) and promoters (FDR < 0.05, 5,458 promoters, **Supp Fig 1B**), followed by TIL-related sites and promoters, with the HBV-driven signatures having the fewest sites or promoters (**Fig 2B, Supp Fig 1B**). In all global DM signatures, there is a bias towards hypomethylation (**Fig 2B, Supp Fig 1B**). We identified overlapping significant DM sites among global and within patient regional mITH DM signatures to examine the contribution of general cancer or immune specific methylation patterns to regional methylation gradients. We observed unexpectedly little overlap in DM sites among global and regional comparisons (**Fig 2D**) and among mITH DM profiles between patients. Instead DM overlaps came from global comparisons, intra-patient overlaps, with also some individual patient regions overlapping with global DM sites. This suggests that mITH DM profiles are not only unique on a per-patient basis but may also be uniquely independent of general tumor-or immune-driven transcriptional regulation programs.

### Enhancer overlap with differential methylation signatures

We overlapped DM sites with known enhancer regions reported in the HACER database and identified sites residing in putative promoter regions defined as within 2Kbp downstream or 1Kbp upstream of a transcription start site (TSS). While most DM sites fall into promoter regions, sites whose genomic coordinates overlap with a known enhancer region and a putative gene promoter are termed ‘dual-overlap’, regions. We find an aberrant, global TIL-associated bias towards hypermethylation in enhancers, and a less pronounced hypermethylation in dual-overlap sites (**Supp Fig 2A)**. This hypermethylation bias in enhancers is preserved in intra-patient, regional-relative mITH DM sites (**Supp Fig 2B**). Hypermethylation occurring at enhancers but not promoters in samples with a bias for TIL may indicate the activation of TIL associated genes due to promoter hypermethylation often silencing transcription while hypermethylation of enhancers can also be activating. This phenomenon of immune infiltration associated with enhancer, not promoter, DMR has been reported in squamous lung cancer,^15^ and conversely with recurrent enhancer hypomethylation associated with poor prognosis in HCC.^16^

### mITH signature is not a simple recapitulation of transcriptomic profiling

Since ITH is a complex manifestation of cancer evolution, we combined DM promoters with differentially expressed (DE) genes from RNAseq data to gain an integrative view of the data.^3^ DM promoters and DE genes had zero overlap for 32 out of 39 regional comparisons across 9 patients. A handful of regional comparisons, which belong to patients 2, 3, 4, and 5, had low overlap relative to the DNA methylation (max = 34.06%, median = 17.07%) and to the RNA (max = 10%, median = 1.590%) (**Fig 3B**). Sample sizes from gene overlap only allowed us to correlate DNA methylation and gene expression log-fold changes in three contrasts, with no significant correlation in two samples (H4.c and H4.a&c) and a moderate negative correlation in the third (H3.a; Spearman rho = −0.409, P = 4.5 e-03) (**Fig 3B)**. Taken together with existing literature,^11,12^ these observations suggest that the mITH described by DM profiling offers a unique signal that may not be captured by gene expression information alone.

### Tumoral hyper-aging and inter-regional heterogeneity predicted by methylation clock

To quantify how mITH relates to tumor clonal evolution, we calculated the predicted methylation age of the tissue across tumor samples and adjacent normal tissue using methylation clocks.^11^ While only 12 CpG sites of the 353 used to inform Horvath’s model had reasonable coverage in the RRBS sequencing data (**Fig 1C**), the promoter-aggregated site analysis provided coverage overlap at the gene level of 240 of the 353 genes from the methylation clock mode (**Fig 1C**). This allowed us to predict tissue methylation age across samples at the gene level. To visualize how mITH might be driving downstream differences in clonal evolution, we compared the relative tissue methylation age across tumor and normal for patients 2, 4, 8, 9 and 10, each of which had three or more regionally sampled tumor sites and sampled adjacent normal tissue (**Fig 4A**). While for all but patient 8 we found wide variance (variance ranges from 3.66e-03 to 1.34e-02 vs 5.63e-04 for patient 8) in the predicted methylation ages across regional samples within tumor, the average relative tumoral methylation age was more advanced than the predicted methylation age of the adjacent normal liver tissue for all patients. Tumoral relative hyper-aging was greatest in patient 8, where average tumoral methylation age was 1.35 times greater than that of the average predicted age of the adjacent liver tissue (**Fig 4B**), with a median tumoral hyper-aging factor of 0.17x across patients. Tumor-normal blood relative methylation age ratios calculated for two of three additional, previously reported, multi-regionally samples HCC patients (GSE83691) validated this trend, though the remaining patient had a significant hyper-aging signature of the adjacent normal tissue compared to the tumor (**Supp Fig. 3**).^4^

**Figure 4.**
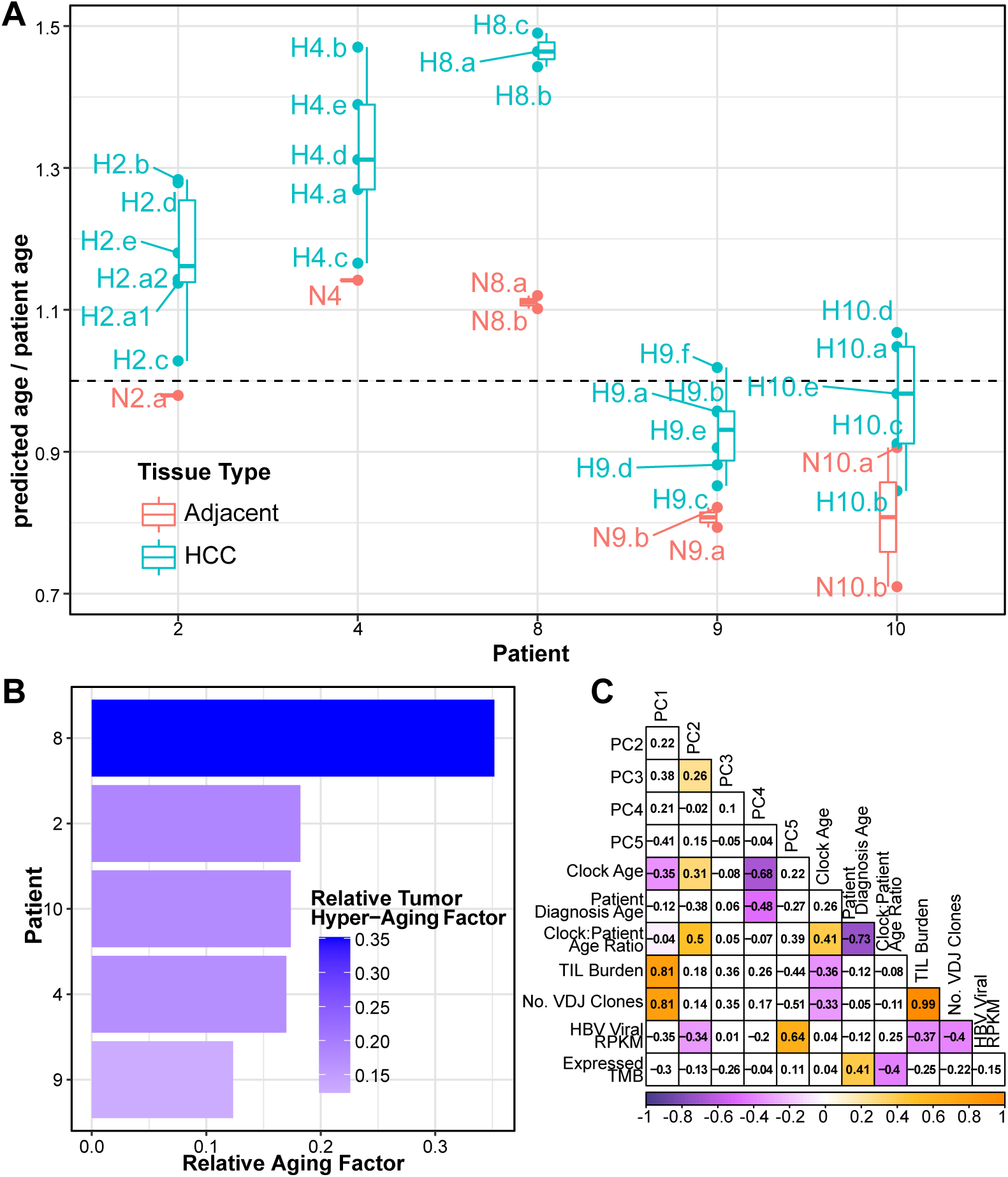
Tumor methylation aging relative to patient age. A) Ratio of predicted tumor methylation age to patient age across regionally sampled patients. B) Hyper-aging relative to tumor methylation age in adjacent normal explains patient 8 outlier status through accelerated tumoral hyper-aging. C) Correlation heatmap of MSSM cohort sample clinical, phenotype, and PCA with methylation clock age and relative age factors across promoter-aggregated regions.

The ratio of predicted sample methylation ages to true patient age was used to determine a relative tissue DNA methylation aging factor. This ratio was significantly positively correlated with the second principal component of a promoter-aggregated site methylation PCA across regional samples (**Fig 4C**; Spearman rho = 0.5, P = 0.002), with the tumoral samples from patient 8 demonstrating the largest hyper-aging effect compared to the true patient age. Predicted methylation age (“Clock age”) was also significantly correlated with PC2, but to a lesser degree (**Fig 4C**; Spearman rho=0.31, P = 0.001). Neither measure correlated with other sample clinical covariates. We also observed a negative correlation between predicted methylation clock age and VDJ read burden (**Fig 4C**; Spearman rho = −0.36, P = 2.52e-04). These data indicate that tumoral hyper-aging is a potentially useful quantitative proxy for mITH.

### Tumoral hyper-aging associates with reduced tumor clonality, decreased tumor fitness, and “hotter”, less clonal tumor microenvironment

We applied our methylation-age prediction analysis to 366 single biopsy HCC samples from the 450k methylation array TCGA-LIHC cohort to validate our observation of tumor hyper-aging and establish the association with patient survival and known clinical biomarkers for immune activity such as TIL and tumor mutational burden (TMB). We observed advanced relative tumor methylation-age in 260/366 (71%) HCC patients in the cohort, with an average methylation aging factor of 1.22 and a median methylation aging factor of 1.15 (ratio of predicted tumoral methylation age to patient diagnosis age; **Fig 5A**). Kaplan-Meier survival estimates were calculated for patients organized by tumor methylation clock age into “old” and “young” groups, thresholded by the median tumor aging factor (1.15). Patients with “old” methylation-aged tumors saw significant survival benefit over patients with “young” tumors (HR=0.667, p=0.037; **Fig 5B**).

**Figure 5.**
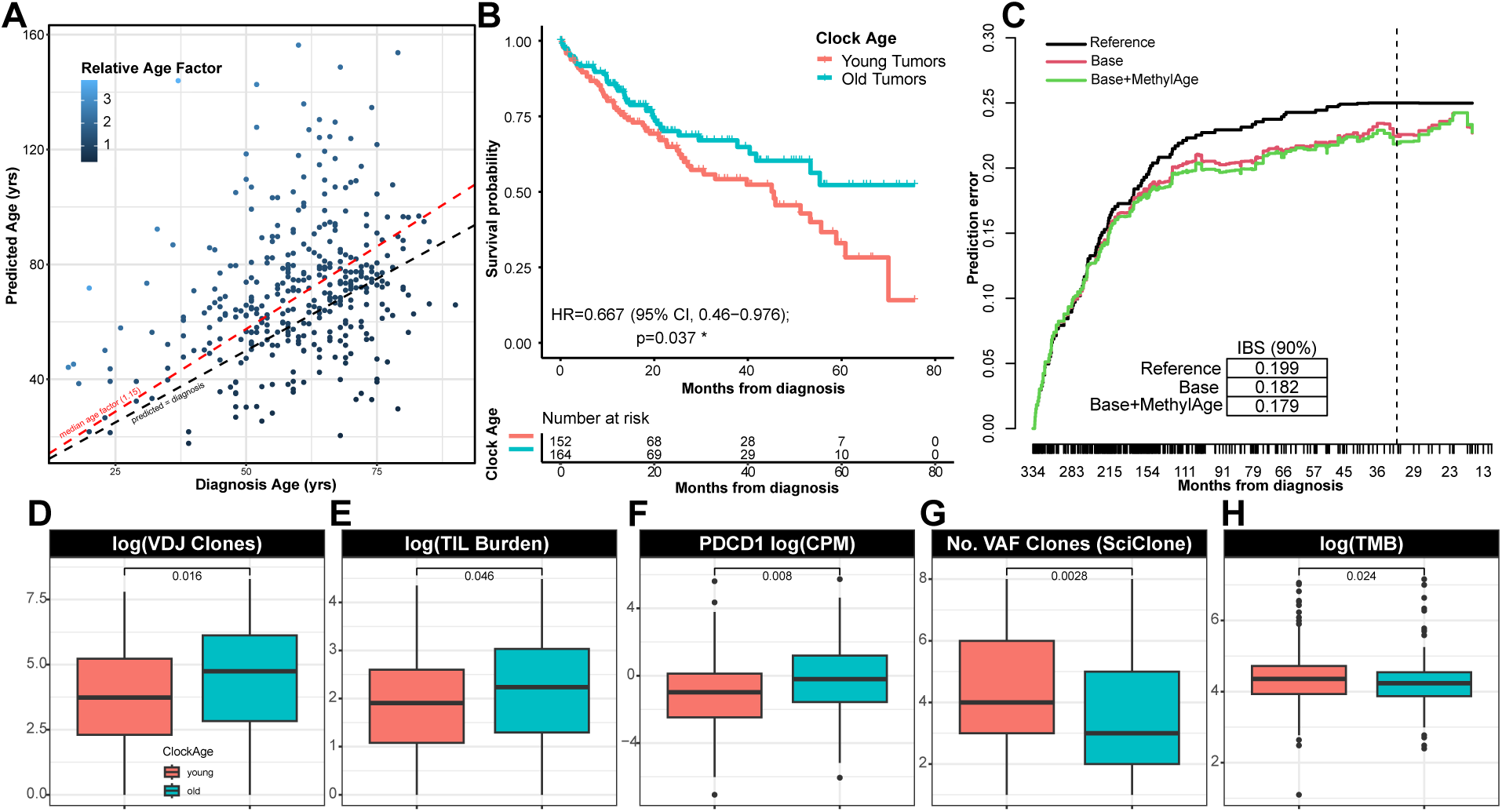
TCGA validation of tumor-methylation age and hyper-aging. A) Age factor landscape of HCC tumors from TCGA LIHC. B) Kaplan-Meier analysis illustrates the survival benefit of “old” tumors with relative hyper-aging signature. C) Comparison of prediction error across cph models. A survival model including the predicted methylation age factor outperforms the model using only covariates of clinical tumor stage, sex, and TMB, across 90% of patient events. D) LIHC HCC relative methylation age factor tracks with known clinical markers including VDJ clone count, E) TIL burden, F) PD-1 expression from RNA-seq G) number of tumor subclones estimated via SciClone, and H) tumor mutation burden (total number of somatic mutations).

To investigate whether the relative tumor methylation age remains a significant predictor of patient survival after regressing out known clinical covariates, we constructed a multivariable Cox proportional hazard (cph) model using patient TMB, sex, tumor stage, and relative tumor age factor (**Supp Table 5**). A regularized cubic spline was applied to the relative age factor to test for non-linear effects on survival. We found relative tumor methylation age to have a significant positive effect on patient survival (p=0.049) (**Supp Table 5**), with no evidence for a significant non-linear effect. To demonstrate the improvement in survival prediction error using relative tumor methylation age, we compared the time dependent integrated Brier scores for nested cph models using the clinical covariates with and without the relative methylation age factor (**Fig 5C**). Comparing the resulting prediction error curves under bootstrap resampling (see Methods) we found that inclusion of relative methylation age factor reduces survival prediction error compared to the model that does not include the measure (**Fig 5C, inset table**). Computing Brier scores under cross-validation indicates these results are robust and will favorably generalize to other datasets.

As we found advanced relative tumor DNA methylation age associated with improved patient survival (decreased tumor fitness) in the TCGA, we investigated its association with other key tumoral and immune molecular features. Performing VDJ-deconvolution of RNA sequencing data from TCGA HCC samples matched with 450k array data,^17^ we observed TIL burden and immune clonality to be significantly higher amongst “old” relative tumor methylation age HCCs compared to “young” tumors (p=0.016, p=0.046; **Fig 5D-E**), and PD-1 receptor expression typically associated with T cell influx was similarly significantly upregulated in “old” tumors (p<0.01; **Fig 5F**). Interestingly, using the TCGA HCC DNA-sequencing data to determine and cluster somatic mutations into clones, “old” HCCs were also found to have significantly fewer tumor clones compared to “young” tumors (p<0.01; **Fig 5G**), and tended to have lower TMBs (p=0.024, **Fig 5H**). These results relate mITH by way of tumoral hyper-aging to reduced clonal complexity, hotter and more clonal TIL burden, and decreased tumor fitness, in a much larger.

## DISCUSSION

DNA methylation alterations in cancer are often difficult to systematically interpret outside of well-studied examples such as promoter hypermethylation of tumor suppressor genes, hypo-methylation of repeat-rich regions. This is true in part because of poor correlation to expression-based characterizations of DNA-methylation changes. This translates into limited evidence on the contribution of DNA methylation to ITH. In this work we provide the most comprehensive analysis of epigenetic ITH in HCC reported to date. Using well-known epigenetic clocks, we show that tumoral hyper-aging, which may be biologically related to a higher mitotic rate in tumor cells, is a useful, novel metric for describing tumor evolution and mITH in both the multi-regional and single-biopsy context. The chronological alteration of epigenetic profiles via progressive DNA methylation has been extensively established in several normal tissues,^18^ however its utility as a proxy of mITH for malignancies has yet to be fully explored. From the point of view of multi-regional sampling, we show that mITH effectively describes differential epigenetic aging for spatially distal regions in HCC relative to the adjacent normal aging rate. In other words, different parts of the same tumor are epigenetically aging at different rates, and on average these rates are all higher than that of adjacent non-HCC liver tissue. Even from the point of view of single-biopsy data such as the TCGA LIHC cohort, we show that, on average, even a single HCC biopsy is older than the diagnosis age of the patient. In our previous work,^3^ we established that a key facet of ITH was immune-editing resulting from tumor-immune interactions which can effectively constrain tumoral evolution.

Regional differential methylation underlying hyper-aging tends to implicate promoter regions, with some overlapping enhancers, belonging to genes which are not differentially expressed in those same regional comparisons reinforcing the importance of mITH in addition to transcriptomics. Moreover, although some mITH sites were shared between individual patients’ signatures and global contrasts such as HCC status or TIL burden, there remains a high degree of patient specificity in mITH patterning and an absence of significant inter-patient mITH site overlap. This indicates highly specific epigenetic reprogramming between patients and can characterize tumor evolution and behavior. Still, patients with marked mITH, such as patients 4 and 5, had strong enrichment in DM signatures associated with TIL burden and HCC vs. adjacent normal liver. This contrasts with patients with minimal mITH, such as patients 7 and 10, who were significantly under-enriched in these contrasts. This underscores that mITH reflects an orthogonal axis of tumoral evolution that is not well captured in a single biopsy context but can be captured in as few as two biopsies (patient 3). This phenomenon of mITH and its effects on tumor clonal evolution can be further assessed through the relative age of the tumor in regions with hyper-aged, “old”, tumors had reduced clonal burden and higher TIL burden. mITH also provides a rich feature space from which to mine potential biomarkers of tumoral evolution and treatment response in HCC. Given the relative granularity of eRRBS data compared to array-based DNA methylation data, these spatial multi-regional mITH data add several previously uncharacterized mITH-specific loci for potential downstream prioritization in other applications, including liquid biopsies.

As of now, there are very few studies that evaluate mITH in cancer. A comprehensive characterization of mITH in lung cancer found that ITH mapping to tumor suppressors was lower than that of oncogenes. The authors suggested a greater selection pressure in these regions with lower methylation ITH.^19^ Another very recent publication in non-small cell lung cancer identified a 25-fold increase in interpatient heterogeneity in tumors as compared to normal samples, as well as inter-patient methylation heterogeneity being greater than mITH.^20^ In HCC, there have been two studies using multiregional sampling and array-based DNA methylation to assess mITH.^4,8^ Unlike ours, the first study^4^ did not include paired gene expression samples but confirmed significant mITH with prognostic value. The second study, focused on driver events of HCC evolution, detects higher levels of hypermethylation in curated epigenetic driver events at CpG islands and promoter regions in addition to the detection of shared and distinct methylation patterns.^8^ Some limitations of our study include a relatively small sample size (despite being among the largest HCC cohorts analyzed for mITH), limited external validation due to the lack of publicly available multi-regional omics datasets, and the absence of a robustly trained epigenetic clock specific for HCC.

This work has characterized an additional layer of molecular intra-tumoral heterogeneity, finding that significant regional differential DNA methylation patterns in patient HCC tumors are unique and poorly recapitulated in standard pooled or single-biopsy profiles. We also report on accelerated epigenetic aging on a regional basis within tumors which serves as a novel proxy measure for ITH. These data can provide new insights into mechanisms underlying ITH, as well as a potential feature space for discovery of novel biomarkers.

## METHODS

### Sample collection

Patient recruitment and sample collection was performed as previously described.^3^ Briefly, patients were enrolled in the study at Icahn School of Medicine at Mount Sinai (ISMMS) and provided informed consent for tissue biobanking. The study was approved by the Mount Sinai IRB (IRB# HS-14-01011) and samples were provided by the ISMMS Tissue Biorepository (IRB# HS-10-00135). All patients had early-stage hepatocellular carcinoma (HCC) per AASLD guidelines and were treatment-naïve prior to surgical resection.^21^ Frozen tissue samples from the same tumor nodule were collected allowing for at least 1 cm of distance between each other. Samples were selected from areas without macroscopic evidence of necrosis or hemorrhage.

### RRBS sequencing and processing

DNA was extracted using the DNeasy blood and tissue kit (Qiagen) from 30mg tissue following manufacturers protocol. RRBS sequencing was performed by the Epigenomics Core facility at Weill Cornell Medical College using an in-house developed protocol consisting of restriction enzyme digestion for enrichment of CpG sites, NGS library construction and bisulfite conversion of cytosines.^14^ 50bp single-read libraries were sequenced using the Illumina HiSeq 2500 platform in high output mode.

Raw RRBS reads were trimmed using trim galore (v0.5.0) with the ‘-rrbs’ flag enabled, and quality control metrics were compiled using fastqc (v0.5.0). Reads were then aligned to the hg38 UCSC reference genome using bismark (v0.22.3), bowtie2 (v2.4.1), and samtools (v1.11) with the options to retain unmapped and ambiguously mapped reads enabled. A count matrix with methylated and unmethylated counts for each sample was created using bismark’s methylation extractor tool (v0.22.3).

### Differential methylation analysis

Differential methylation signatures were computed with edgeR following previously established methodology outlined in Chen et al.^22^ We used the generalized linear model framework in edgeR because it allows for flexibility and robustness in analyzing complex experimental designs while being able to account for technical and biological covariates. Briefly, this approach works by adding extra columns to the design matrix to account for read coverage at each site or promoter, therefore allowing for statistical analyses that mirror those typically used for RNA-seq.^22^ This analysis was performed in two settings: at individual CpG island sites (i.e. site-based), and at the promoter level by aggregating counts falling within the same promoter regions (i.e. promoter-based).

In the site-based analysis, sites with a minimum coverage of 10 reads across all samples were retained. In the promoter-based analysis, counts for sites that fall within the same promoter region were summed the nearestTSS function in the edgeR package.^22,23^ These regions were defined as within 2kb upstream or 1kb downstream of the nearest transcription start site annotation as previously described.^22,23^ Promoters with at least 10 aggregated reads across all samples were retained for further analysis.

Site-based and promoter-based DM signatures were computed on a within-patient, regional-relative basis, and on a global basis. In all analyses, sites, or promoters from mitochondrial, sex, and unannotated chromosomes were removed from analysis. Total library size for all samples was set as the sum of methylated and unmethylated counts. All dispersion estimates assumed no mean-variance trend.

#### Regional-relative DM signatures

Within-patient regionally-relative DM signatures were computed using pooled sample comparisons in an effort to directly sample epigenetic ITH while compensating for lack of statistical power on a per-patient basis. Briefly, a generalized linear model using edgeR’s RRBS pipeline was created for each of the 39 within-patient comparisons across all 9 patients.^22^ HBV viral expression, measured in RPKM, and normalized VDJ read burden were determined via RNA sequencing as previously described and were subsequently included as covariates in the model.^3,22,23^ For each comparison, a separate, paired design matrix that accounts for methylation status in the counts matrix was used. Differential methylation was assessed using a likelihood ratio test via the edgeR package.

#### Global DM signatures

Differential methylation signatures using all pooled tumor and adjacent samples over all patients were computed with generalized linear models using edgeR’s RRBS pipeline. The design matrix accounted for HBV viral expression and normalized VDJ read burden determined as previously described, and multiple samples from the same patient. Differential methylation along three major axes were assessed with a likelihood ratio test: normal vs tumor, VDJ read burden, and HBV viral expression.

### RNA-seq differential expression analysis and integration with DM promoters

To compare expression-based ITH with DM promoter-based mITH, matched RNA sequencing for the MSSM cohort was obtained and processed as previously described.^3^ To ensure a fair comparison between the RNAseq and the RRBS, generalized linear models for DE analysis were generated using the edgeR pipeline with using the same covariates as that used in the DM analysis where reasonably applicable.^23^ DE testing was performed using a likelihood ratio test in edgeR for the same regional-relative and global comparisons used in DM promoter testing as described above.

### Enhancer analysis

Annotations for known enhancers were acquired from the HACER database.^24^ DM Sites were overlapped with the enhancer coordinates and annotated accordingly. DM directional bias in enhancer-residing sites and promoter-residing sites was statistically compared using a Wilcoxon test in R.

### Methylation age prediction

Methylation based tumor age was calculated for the HCC regional samples using the CpG-site based age clock described by Horvath et al.^11^ As site-level coverage of the 353 CpG-sites used to determine methylation age was low in the RRBS data, encompassing genes for the Horvath CpG-sites were identified and beta methylation normalized values for overlapping genes from the promoter-based analysis were used as proxy. Methylation age was only computed for patients with regional sampling of at least one or more normal sites and two or more tumor sites (patients 2, 4, 8, 9, and 10), such that age predictions could be compared intratumorally and between tissue types. Patient age was transformed via logit function as described previously.^11^ Median computed tumor and normal regional sampling ages for each patient were contrasted to calculate the patient normalized relative aging factor ratio.

### Survival Analysis

Kaplan-Meier curves and risk tables grouping patients along the median tumor methylation age factor into “old” and “young” cohorts were created using the survminer R package.^25^ Patients with missing survival times or model feature data were removed before modeling. The log-rank test was used to evaluate the difference in survival outcomes between groups, with the significance and confidence intervals reported for each comparison.

Multivariate cox proportional hazard models using tumor methylation age, tumor clinical stage, sex, and patient tumor mutation burden (TMB), were constructed using the ‘cph’ function from the rms package.^26^ The relative predictive power amongst survival models and to the reference model was assessed by plotting prediction error curves with the pec package in R. Integrated Breier scores for each model were computed with .632+ bootstrap resampling across 100 iterations (B=100) and compared, with the weights of each score corresponding to the inverse likelihood of being censored, with censoring times for each iteration estimated using the Kaplan-Meier estimator. Brier scores for each model were compared at 75 months with significance by KS-test calculated and included in the inset figure tables, corresponding to 90% of patient events.

### Validation of clock-associated tumoral hyper-aging and outcome analysis in external datasets

Two additional Illumina 450k methylation array HCC datasets were used for validation of methylation-based tumor aging and patient survival findings. To demonstrate the tumor hypermethylation and regional methylation patterns seen in the main dataset, we retrieved beta-normalized illumina 450k methylation array data from the GEO database project GSE83691 for 17 samples from multi-regionally sampled HCC liver biopsies for three patients (*HCC8010, HCC6952*, and *HCC8257*) was retrieved.^4^ These three patients were selected due to the availability of matched normal data from circulating blood. Methylation based age predictions were made for each sample and relative aging factor ratios between median tumor ages and normal age were calculated for each patient.

To demonstrate the effects of tumor methylation age on patient survival, Illumina 450k methylation array data for 377 tumor samples from the TCGA liver cancer cohort (LIHC) was retrieved from the GDC portal.^17^ Clinical phenotype and survival data for these patients was also retrieved. TMB information was available for 366 samples, and tumor clonality was assessed using SciClone in 192 samples where appropriate matched mutation data was available.^27^ For 188 samples where matched RNA-seq data was available, the estimated VDJ read burden and TIL clonality was assessed using MixCR, and logCPM expression values for PDCD1 were calculated.^28^ Normalized methylation beta values were calculated for each site, and tumor methylation-based ages were predicted using Horvath’s clock. Relative tumoral age factors were calculated for each patient using the ratio of the predicted tumor methylation age to the patient’s age at diagnosis. Patients were separated into ‘old’ and ‘young’ tumor groups based on median predicted age factor, and survival analyses were carried out as described above. Tumor clonality, VDJ burden, TMB, and PDCD1 expression was statistically compared between tumor groups using a Wilcoxon rank-sum test in R.

## Data Availability

All data and code available upon reasonable request and will be available on github upon publication.

## DATA AND CODE AVAILABILITY

RNA sequencing data is deposited in ArrayExpress under the accession E-MTAB-5905. Reduced-representation sequencing data is deposited in SRA under the accession PRJNA662974. Sequencing data from patient 5 are not deposited due to lack of patient-specific deposition consent. TCGA-LIHC data were generated and made available by the TCGA Research Network (https://www.cancer.gov/tcga) and were downloaded from the National Cancer Institute’s GDC Data Portal (https://portal.gdc.cancer.gov/). <u>Data and code will be available on GitHub upon</u> <u>publication.</u>

## ETHICS STATEMENT

All tumor samples used in this study were collected from consented patients enrolled in the Icahn School of Medicine at Mount Sinai institutional IRB-approved cancer biorepository tissue procurement protocol.

## FUNDING

This work was funded by the Genetics and Genomics department, Icahn School of Medicine at Mount Sinai, as well as the Icahn Institute for Data Science and Genomic Technology. AV is supported by the NIH (1U01CA283931-01)

## AUTHOR CONTRIBUTIONS

B.L. and A.V. conceived the study concept and experimental design. PR and AB analyzed the data under supervision from BL. PR, AB, DCC, BL, and AV wrote the manuscript. A.J.C collected and prepared samples for RRBS sequencing.

## ACKNOWLEDGEMENTS

The authors thank the office of Scientific Computing and the Genomics Core Facility at the Icahn School of Medicine at Mount Sinai (ISMMS) for providing computational resources and staff expertise, as well as the ISMMS Tisch Cancer Institute Biorepository for providing the samples.

## DISCLOSURES

AV has received consulting fees from FirstWorld, Pioneering Medicine and Genentech; advisory board fees from BMS, Roche, Astra Zeneca, Eisai, and NGM Pharmaceuticals; and research support from Eisai. He has stock options from Espervita and Atzeyo. He is listed as an inventor on a patent related to early detection of HCC (PCT/US20/61441).

---

**Supplementary Figure 1.**
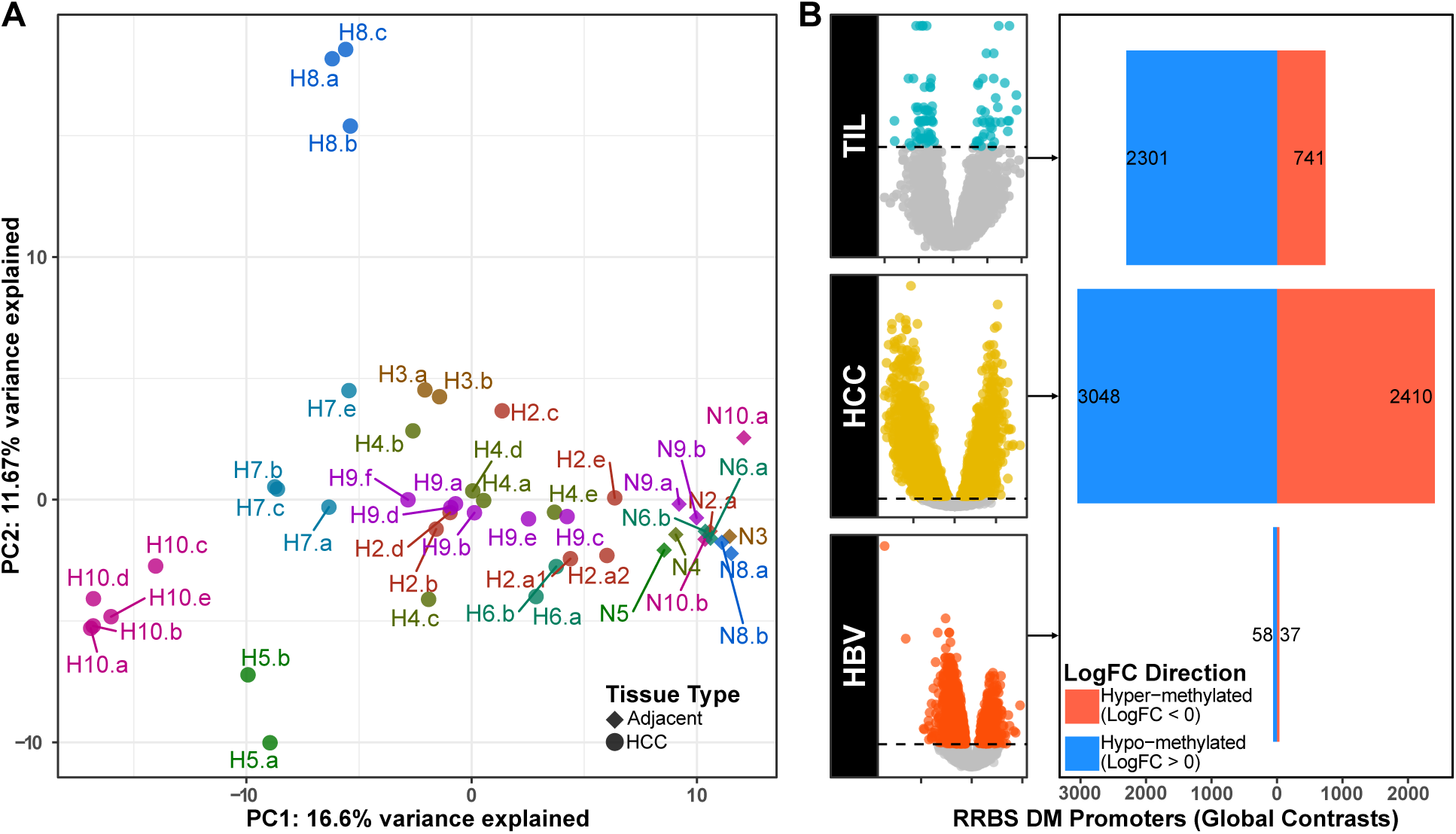
Regional promoter methylation and global promoter contrasts. A) PCA of promoter aggregated methylation beta matrix. B) RRBS DM promoters for global contrasts of (top to bottom) TIL, Tumor vs Adjacent, and HBV.

**Supplementary Figure 2.**
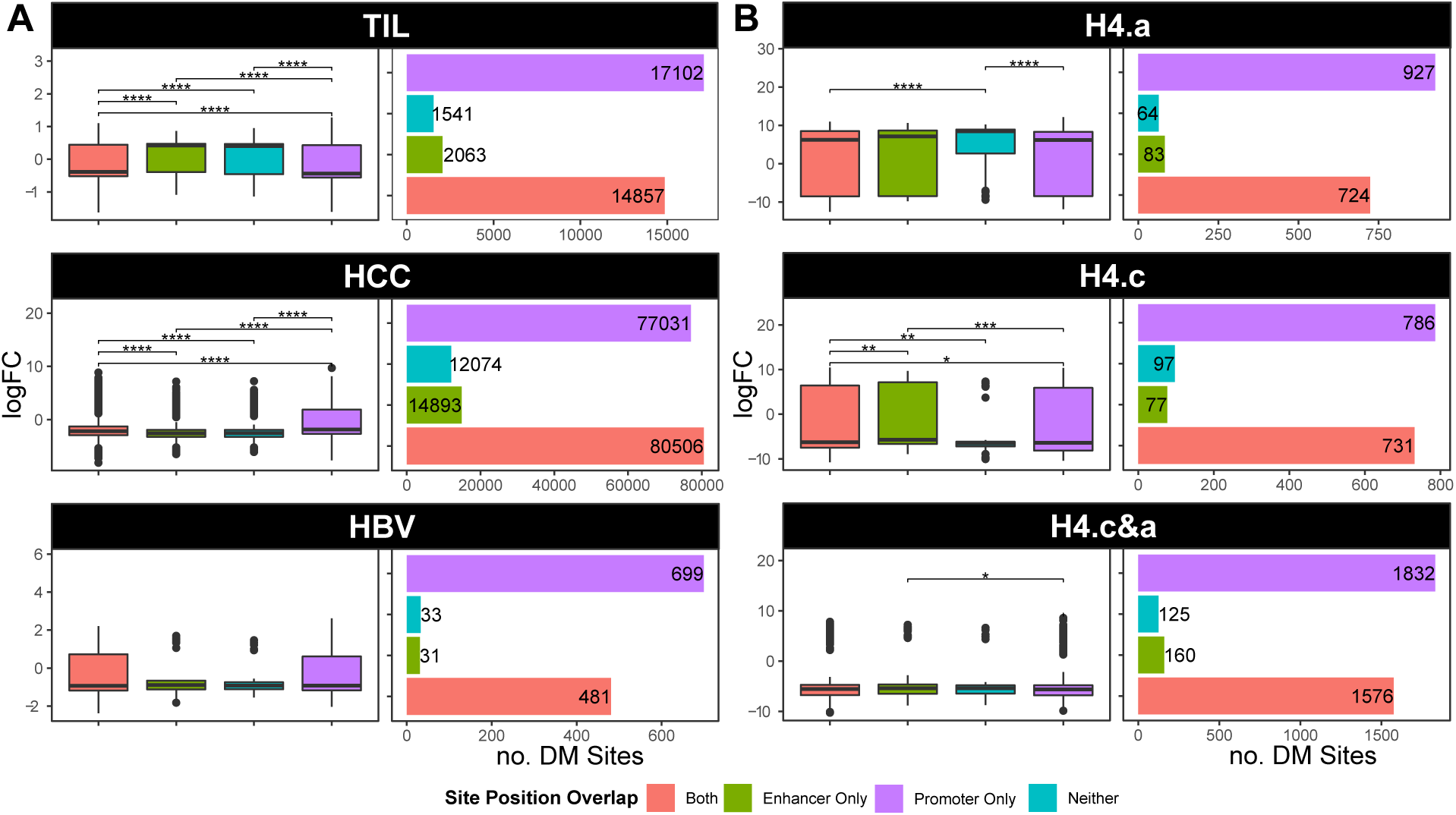
Methylation change comparisons across DM regions covering promoter, enhancer, both, or neither feature loci for A) global contrasts (top to bottom) TIL, Tumor vs Adjacent, and HBV; B) regional contrasts across HCC in patient 4.

**Supplementary Figure 3.**
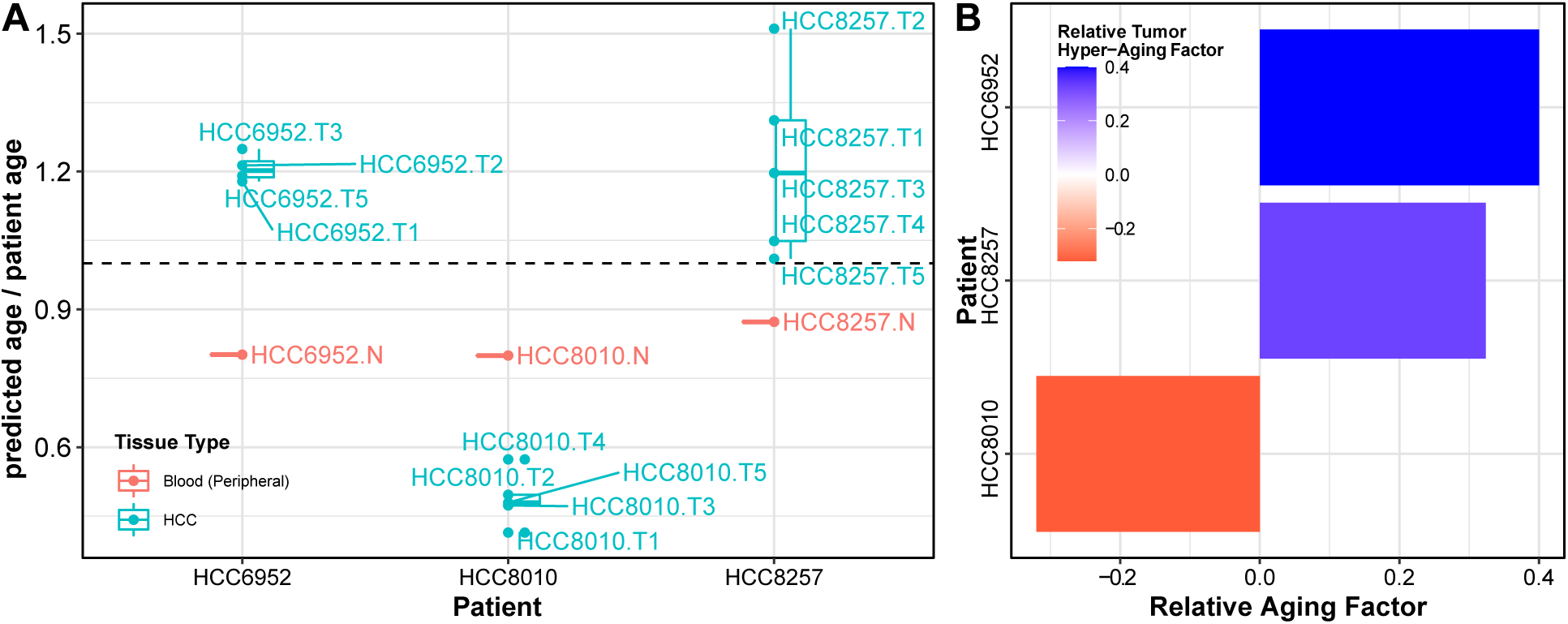
Tumoral hyper-aging validation via GSE83691.^4^. A) Site-by-site methylation predicted age to patient age ratios across three multi-regionally sampled HCCs. B) Tumoral methylation hyper-aging observed in two tumors, and one with hypo-aged tumor.

## Notes

### Competing Interest Statement

The authors have declared no competing interest.

### Author Declarations

The study was reviewed and approved by the Icahn School of Medicine at Mount Sinai IRB in 2014, with approval number HS#14-01011

### Summary of Updates

Manuscript has been revised to improve clarity, readability, and to update the figures to include additional data from the text.

## REFERENCES

1. Bray, F., et al., Global cancer statistics 2018: GLOBOCAN estimates of incidence and mortality worldwide for 36 cancers in 185 countries. CA Cancer J Clin, 2018. 68(6): p. 394–424.

2. Villanueva, A., Hepatocellular Carcinoma. N Engl J Med, 2019. 380(15): p. 1450–1462.

3. Losic, B., et al., Intratumoral heterogeneity and clonal evolution in liver cancer. Nat Commun, 2020. 11(1): p. 291.

4. Lin, D.C., et al., Genomic and Epigenomic Heterogeneity of Hepatocellular Carcinoma. Cancer Res, 2017. 77(9): p. 2255–2265.

5. Zhai, W., et al., The spatial organization of intra-tumour heterogeneity and evolutionary trajectories of metastases in hepatocellular carcinoma. Nat Commun, 2017. 8: p. 4565.

6. Craig, A.J., et al., Tumour evolution in hepatocellular carcinoma. Nat Rev Gastroenterol Hepatol, 2020. 17(3): p. 139–152.

7. Friemel, J., et al., Intratumor heterogeneity in hepatocellular carcinoma. Clin Cancer Res, 2015. 21(8): p. 1951–61.

8. Ding, X., et al., Genomic and Epigenomic Features of Primary and Recurrent Hepatocellular Carcinomas. Gastroenterology, 2019. 157(6): p. 1630–1645 e6.

9. Villanueva, A., et al., DNA methylation-based prognosis and epidrivers in hepatocellular carcinoma. Hepatology, 2015. 61(6): p. 1945–56.

10. Tischoff, I. and A. Tannapfe, DNA methylation in hepatocellular carcinoma. World J Gastroenterol, 2008. 14(11): p. 1741–8.

11. Horvath, S., DNA methylation age of human tissues and cell types. Genome Biol, 2013. 14(10): p. R115.

12. Horvath, S. and K. Raj, DNA methylation-based biomarkers and the epigenetic clock theory of ageing. Nat Rev Genet, 2018. 19(6): p. 371–384.

13. Zhu, T., et al., CancerClock: A DNA Methylation Age Predictor to Identify and Characterize Aging Clock in Pan-Cancer. Front Bioeng Biotechnol, 2019. 7: p. 388.

14. Garrett-Bakelman, F.E., et al., Enhanced reduced representation bisulfite sequencing for assessment of DNA methylation at base pair resolution. J Vis Exp, 2015(96): p. e52246.

15. Cho, J.W., et al., The importance of enhancer methylation for epigenetic regulation of tumorigenesis in squamous lung cancer. Exp Mol Med, 2022. 54(1): p. 12–22.

16. Xiong, L., et al., Aberrant enhancer hypomethylation contributes to hepatic carcinogenesis through global transcriptional reprogramming. Nat Commun, 2019. 10(1): p. 335.

17. Cancer Genome Atlas Research Network. Electronic address, w.b.e. and N. Cancer Genome Atlas Research, Comprehensive and Integrative Genomic Characterization of Hepatocellular Carcinoma. Cell, 2017. 169(7): p. 1327–1341 e23.

18. Bell, C.G., et al., DNA methylation aging clocks: challenges and recommendations. Genome Biol, 2019. 20(1): p. 249.

19. Hua, X., et al., Genetic and epigenetic intratumor heterogeneity impacts prognosis of lung adenocarcinoma. Nat Commun, 2020. 11(1): p. 2459.

20. Gimeno-Valiente, F., et al., DNA methylation cooperates with genomic alterations during non-small cell lung cancer evolution. Nat Genet, 2025. 57(9): p. 2226–2237.

21. European Association For The Study Of The, L., R. European Organisation For, and C. Treatment Of, EASL-EORTC clinical practice guidelines: management of hepatocellular carcinoma. J Hepatol, 2012. 56(4): p. 908–43.

22. Chen, Y., et al., Differential methylation analysis of reduced representation bisulfite sequencing experiments using edgeR. F1000Res, 2017. 6: p. 2055.

23. Robinson, M.D., D.J. McCarthy, and G.K. Smyth, edgeR: a Bioconductor package for differential expression analysis of digital gene expression data. Bioinformatics, 2010. 26(1): p. 139–40.

24. Wang, J., et al., HACER: an atlas of human active enhancers to interpret regulatory variants. Nucleic Acids Res, 2019. 47(D1): p. D106–D112.

25. Kassambara, A., Kosinski, M. & Biecek, P., survminer: Drawing Survival Curves using ‘ggplot2’. 2020.

26. Harrell, F.E., rms: Regression Modeling Strategies. 2020.

27. Miller, C.A., et al., SciClone: inferring clonal architecture and tracking the spatial and temporal patterns of tumor evolution. PLoS Comput Biol, 2014. 10(8): p. e1003665.

28. Bolotin, D.A., et al., MiXCR: software for comprehensive adaptive immunity profiling. Nat Methods, 2015. 12(5): p. 380–1.

